# The impact of deprivation and neighbourhood food environments on home food environments, parental feeding practices, and child eating behaviours, food preferences and BMI: The Family Food Experience Study-London

**DOI:** 10.1101/2025.02.07.25321815

**Authors:** Andrea Smith, Alice Kininmonth, Kristiane Tommerup, David Boniface, Chiara Gericke, Tiffany Denning, Carolyn Summerbell, Christina Vogel, Clare Llewellyn

## Abstract

**Purpose:** Childhood obesity inequalities in England persist despite targeted interventions focused on promoting healthy diets and food environments. This study, part of the Family Food Experience Study-London, aimed to investigate the impact of deprivation and neighbourhood food environments on home food environments, parental feeding practices, child eating behaviours, food preferences, and child BMI.

**Methods:** Families (n=728) with primary school-aged children were recruited from four socioeconomically diverse London boroughs in 2022. Data were collected through computer-assisted interviews (30.8% in-person, 69.2% telephone) on home food environment, parental feeding practices, and children’s eating behaviours and food preferences. Deprivation was characterised using a composite measure of family and neighbourhood indicators of socioeconomic position. Neighbourhood food environment exposures were derived from individualised activity spaces. Child BMI was measured objectively. Generalised linear models examined associations between deprivation and neighbourhood food environment with family food-related outcomes, adjusting for school-level clustering, child sex, age and ethnicity.

**Results:** Greater neighbourhood deprivation was significantly associated with more ‘obesogenic’ family food practices, child eating behaviours and child BMI. Deprivation was linked to higher food responsiveness (β=-0.12, p=0.002), emotional overeating (β=-0.11, p <0.001), and increased desire to drink (β=-0.26, p <0.001). Parents in deprived households used more emotional (β=-0.10, p<0.05), instrumental (β=-0.11, p=0.003) and pressuring feeding practices (β=-0.14, p<0.001). Greater deprivation was also associated with a more obesogenic home food environment (β=-0.19, p<0.001) and lower meal structure (β= 0.17, p<0.001). Exposure to less healthy neighbourhood food environments around and between home and school were associated with a more obesogenic home food environment (β=-0.07, p<0.01), but no significant associations were found with feeding practices or child eating behaviours or child BMI.

**Conclusions:** Family deprivation, rather than neighbourhood food environments, is more strongly linked to obesogenic feeding practices, child eating behaviours and child BMI. Policies focusing on improving neighbourhood food environments will likely be most effective if combined with those addressing systemic issues related to deprivation such as welfare policies (e.g. reforms to benefit caps) or targeted subsidies for healthy food. Future research should examine the independent and accumulative impact that environment and household interventions have on childhood obesity inequalities.

## BACKGROUND

In England, the prevalence of childhood obesity in the most deprived neighbourhoods is more than double that in the least deprived areas. These trends are observed among children in the first (age 4-5: 12.9% versus 6.0%) and last (age 10-11: 29.2% versus 13.0%) years of primary school^1^. Stark inequalities in childhood obesity persist despite targeted policies aimed at improving children’s diets nationally and locally^2^. London has long had the highest poverty rate of the United Kingdom (UK) regions^3^ and the prevalence of childhood obesity in London is also one of the highest in England^4^. Tackling inequalities in childhood obesity in the London area is therefore a public health priority., Local Authorities (LAs), however, do not fully understand why the range of food-related policies and interventions in place are not working effectively for all children. Child poverty is a major, modifiable driver of poor health and lifelong inequalities, affecting everything from child mortality and mental health to school readiness and life chances. Without addressing it, other health interventions aimed at reducing obesity are likely to fail^5^. The UK government recommends a whole systems approach to tackling obesity^6^, which considers individuals relationships and behaviours within the wider context in which they live. To support the development of systems approaches to childhood obesity inequalities across London, the Family Food Experiences Study-London (FFES-L) is a large, multi-disciplinary complex systems project which aimed to understand how the context of deprivation prevents families in London from engaging with, and benefiting from, existing local public health interventions focused on healthy eating and drinking. This information will allow LAs to adapt and augment existing interventions to better meet the needs of local families who would benefit most, and effectively narrow the inequalities gap in childhood obesity ^7–9^.

Stark changes to the foodscape over the last few decades, such as increases in the number and density of unhealthy food retail outlets, are thought to have contributed to excess energy intake among children ^8,9^. A more recent systematic review concluded that the number of, and distance to, unhealthy food outlets was associated with greater consumption of fast-foods and higher BMI in children^10^. There is also a strong linear association between fast-food outlet density and neighbourhood-level deprivation in England, implicating the quality of the local food environment to inequalities in childhood obesity^11,12^.

More proximal aspects of the home family environment may also drive inequalities in childhood obesity, given that socioeconomic patterning in weight gain starts to emerge even in the first few months of life^13^. Parents are the ‘gatekeepers’ and decision-makers to young children’s food and drink access. As such, the quality of the home food environment, and the feeding practices and policies that parents adopt, can shape children’s eating behaviours and weight. Both the ‘what’ and the ‘how’ of the early feeding environment matter in this context. Children who live in a home with greater availability of, and access to, healthier foods have a better diet quality^14–16^. At the same time, non-responsive parental feeding practices (using food for non-nutritive purposes, such as to control a child’s behaviour or to soothe their emotions) are generally associated with less desirable eating behaviours and higher weight. Providing structure around mealtimes and eating, and monitoring a child’s food and drink intake, have been linked to more favourable outcomes^17,18^. There is some evidence of socioeconomic patterning in the quality of the home food environment, and in parental feeding practices^19^, but most prior research has focused on maternal education only, rather than capturing the complexity of deprivation that spans neighbourhood, household and individual factors.

Children’s own eating styles are also important in determining what and how much they eat. Children’s appetitive traits (stable tendencies towards food and the opportunity to eat) are well-established risk factors for obesity. ‘Food approach’ traits include a tendency to want to eat in response to negative emotions. If palatable food or drink is easily available in the home or proximal food environment (e.g. corner shop); they are robustly associated with obesity risk in childhood^20^. Greater socioeconomic deprivation has been linked to increases in the expression of ‘food approach’ traits from toddlerhood through to early childhood^21^, but the reasons underpinning this relationship are not clear. Poorer quality proximal (e.g. home) or local (e.g. neighbourhood) food environments are likely to contribute. Additionally, food preferences are crucial modifiable determinants of what children eat^22^, yet the extent to which food likes and dislikes vary with deprivation is largely unknown. Food likes and dislikes may pose important barriers or facilitators to successful uptake of local interventions that focus on healthy food provision highlighting an important evidence gap.

This multidisciplinary study addressed these evidence gaps by integrating different levels of information from families with primary school-aged children living in four London Boroughs. The study aimed to: 1) identify the different domains of family food-related outcomes that vary with deprivation and the neighbourhood food environment; and 2) examine whether the context of deprivation interacts with the local food system to shape family food-related outcomes.

## METHODS

### Study design and sample

A sample of socioeconomically diverse families (n=739) from four London boroughs (Southwark, Haringey, Greenwich, or Croydon) were recruited via primary schools. Data were collected via a comprehensive computer assisted personal interview (CAPI) with one caregiver per household. Families were recruited through a targeted strategy that disseminated invitations to participate in the study via primary schools in two rounds. In the first round (19th April – 4th June 2021), the National Centre for Social Research (NatCen) sampled 28 of 152 schools in the lowest 25% of Income Deprivation Affecting Children Index (IDACI) (least deprived; n=14 schools) and highest 25% (most deprived; n=14 schools), to ensure a socioeconomically diverse sample. IDACI is an index of deprivation used in the UK which measures in a local area the proportion of children (<16y) that live in low-income households ^23^. The first round of recruitment did not achieve the required number of parent opt-ins, so the recruitment strategy was modified to approach n=136 schools with a mid-range IDACI score. The second round (31st August to 31st December 2021) generated a further 30 schools. A flow diagram of the recruitment is shown in **Fig S1**.

Schools received a book token (£300) for sending out study invitations to parents (from Reception to Year 6) via email, school newsletters, or digital flyers on parents’ WhatsApp groups. Parents agreed to participate via an online link where they provided basic sociodemographic information and contact details. Once recruitment had been completed, families with a child in the target age group, who attended a school in an eligible postcode (for Southwark, Haringey, Greenwich or Croydon), were contacted by NatCen to arrange the day/time for the survey interview and to select a home visit or telephone call. All measures were collected during the interview (see **Supplementary materials** for information on measures used in this study; the interview schedule is provided alongside the full dataset). Each interview was approximately 1 hour, with about 8 minutes for anthropometric measurements and set-up. Upon completion, families were offered a £30 gift voucher for their participation.

Prior to data collection, a pilot study was undertaken to test and refine the recruitment and survey (May-June 2021). Details of the pilot feasibility study are provided in the **Supplementary Information**. The main data collection took place between 13^th^ September 2021 and 29^th^ May 2022. The flowchart for recruitment and sampling is shown in **Figure S1.** Data were collected for 739 families whose children were aged between 4-11 years. Of these, 50 families (6.76%) whose home postcodes were outside the four London boroughs, were classified into one of the four boroughs based on their school postcode. Nine families were excluded from analyses because both their home and school postcodes were outside the four boroughs; two families were excluded because they did not complete the interview. This left a final sample of n=728 families for analysis.

### Measures

#### Socioeconomic Position

The primary caregiver provided information about multiple indicators of socioeconomic position (SEP), including: highest maternal educational qualification; current occupation (both parents, if applicable); total annual household income; postcode (for calculating index of multiple deprivation, IMD)^23^; home ownership status; number of bedrooms in the home; number of cars (**Figure 1**). Principal component analysis was used to create a composite score, which incorporated all seven indicators, spanning individual, household, and neighbourhood-level factors to capture the complexity of SEP. Higher scores reflected higher SEP (sample range: 0.94–6.94). Details about the SEP composite score are available elsewhere ^21^.

**Figure 1.**
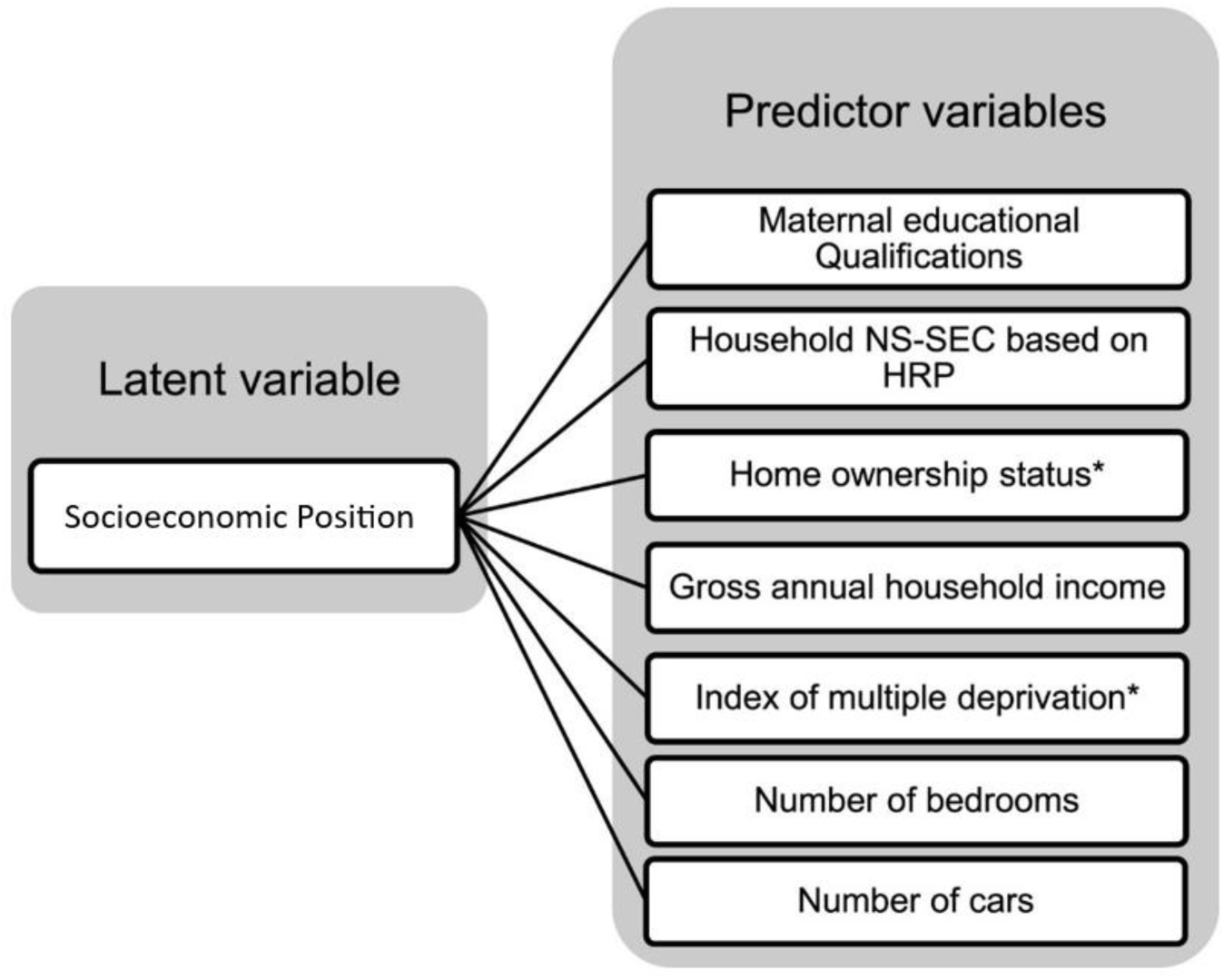
The seven indicators of socioeconomic position that were included in the composite measure of SEP (*item reverse scored). Abbreviations: NS-SEC=The National Statistics Socio-economic classification; HRP=Household reference person.

#### Neighbourhood food environment

The neighbourhood food environment around the home, the school, and the route in between, was characterised for each child using Ordinance Survey Point of Interest (POI) data which specifies the physical location of different types of food outlets. The route between the home and school was calculated using the road network distance between the home and school locations and assigning a mode of travel (walk mode if <3km; drive mode if >3km). The travel mode defined the street/road network, which was used to create travel routes with the shortest road network route between home and school was calculated for each participant. Travel areas were generated around each participant’s home and school location, using a network buffer of 500m. A 50m buffer zone was generated around each roadside of the shortest route between home and school. The buffer zone around the home, school and the route in between is termed the child’s ‘activity space’ (**Figure 2**).

**Figure 1.**
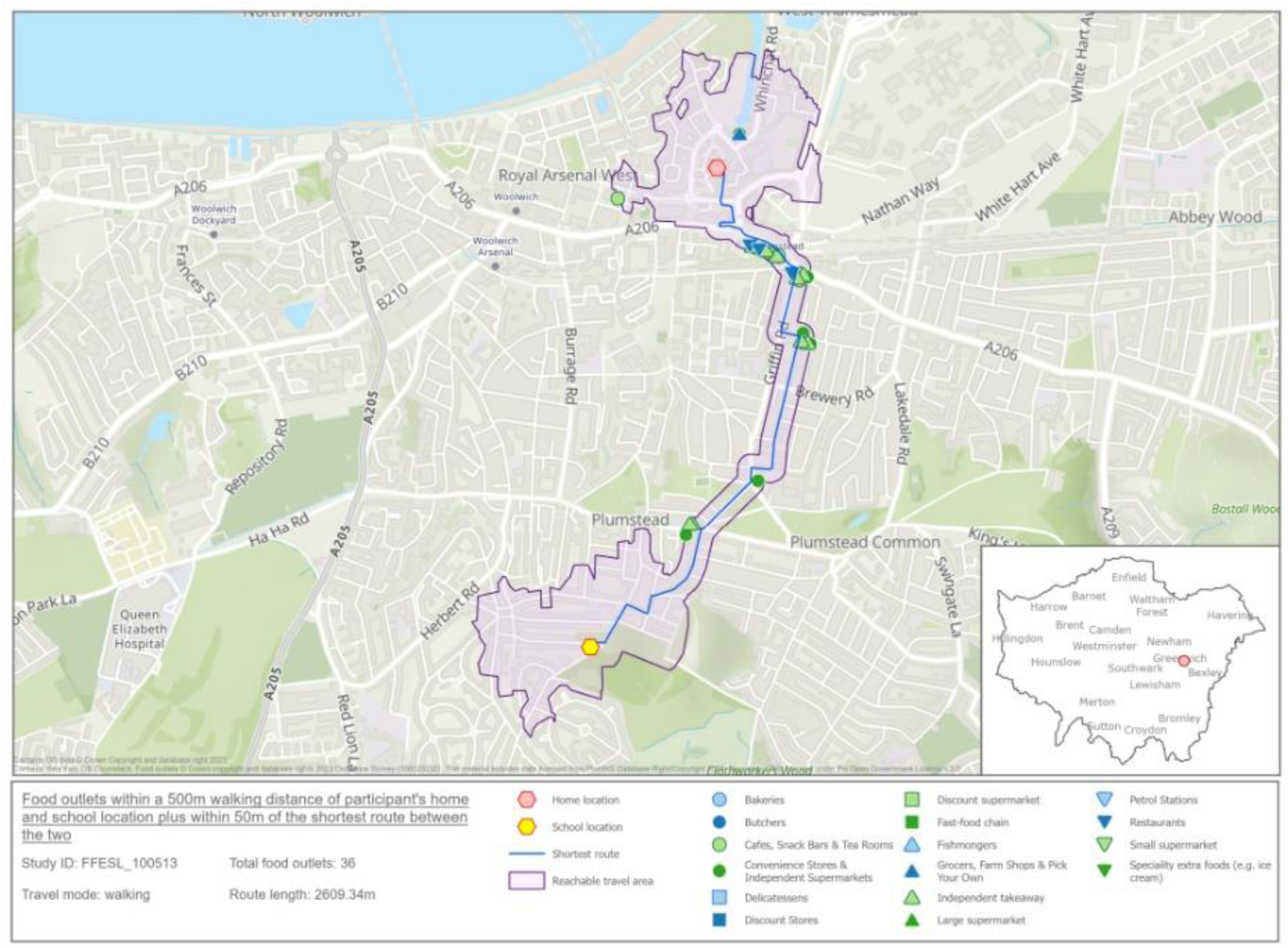
The area given in purple includes 500m walking distance service area generated around the home location and school location and 50m buffer within the route from home to school. The food outlets within this area are shown, see key for more information about the specific food outlets. Image contains OS data *© Crown Copyright and database right 2023 Contains data from OS Zoomstack*

Using a similar approach applied in previous food environment research^24^, a neighbourhood score for each child was calculated that represented both the type and number of food outlets they were exposed to in their activity space. These scores represented both spatial exposure to different types of food outlets and a proxy for the healthfulness of the in-store environments. Scores were calculated by: (i) identifying the number of each type of food outlet within the child’s activity space; and (ii) multiplying the number of each food outlet by a weight describing the relative availability of healthy and unhealthy foods within each type of food outlet. Weights were determined from a previous Delphi study^25^. Higher scores represent a healthier food environment (scores ranged −1349.0 to 142.0). Further information about the scoring is provided in the **Supplementary information**.

#### Home food environment

The home food environment was characterised using a comprehensive measure which assesses physical and social aspects of the home food environment and has been validated in preschool- and school-aged children^14,15^. The ‘obesogenic’ quality of the home food environment was determined by creating a composite score, with a higher total score reflecting ‘higher-risk’ for excess weight gain. Parental feeding practices were removed from the score and examined separately.

#### Parental feeding practices

Nine parental feeding practices (PFPs) were assessed: four nonresponsive PFPs (Instrumental feeding, Emotional feeding, Pressure to eat, Restriction); and five responsive PFPs (Parent control, Monitoring, Encouragement to eat nutritious foods, Modelling, Covert restriction). All items were rated using a five-point Likert scale from ‘never’ (1) to ‘always’ (5), except the restriction scale which was measured on a 7-point Likert scale from not at all (1) to strictly (7). A mean score was calculated for a scale if responses were available for most items within a scale. All measures have been validated in comparable populations, except for the restriction scale^26–29^. Further detail on items and internal scale reliability is available in **Table S4.**

#### Child eating behaviours

Child appetitive traits were assessed using the Children’s Eating Behaviour Questionnaire (CEBQ)^30^. The CEBQ is a 35-item parent-report psychometric measure of eight appetitive traits (seven eating behaviours and one drinking behaviour), rated using a 5-point Likert scale (1=Never to 5=Always). It has been validated using behavioural measures of food intake and has good internal and test-retest reliability^30,31^. The traits can be broadly split into four *food approach* traits and four *food avoidant* traits. Food approach traits characterise a more avid appetite and greater interest in food and include Food Responsiveness, Enjoyment of Food, Emotional Overeating, Desire to Drink. The food avoidant traits characterise a less avid appetite and lower interest in food and include Satiety Responsiveness, Slowness in Eating, Food Fussiness and Emotional Undereating. A mean score was calculated for each subscale if participants completed >half the items for that scale. Further details, including example items and internal scale reliability, can be found in **Table S4**

#### Child food preferences

Parents rated their child’s liking of 75 foods using a 5-point Likert scale, ranging from likes a lot (1), to dislikes a lot (5). Participants could also select ‘never tried’, which was recoded to missing. Responses were reverse coded so that higher scores reflected higher preference for a specific food. Foods were categorised into six groups: fruit, vegetables, protein, dairy, starches and snacks. Further information about the development of the food preference questionnaire has been published elsewhere^32^. Internal scale reliability is available in **Table S4.**

#### Child anthropometric measures

For interviews completed during home visits, child height and weight were measured by the trained NatCen researcher using portable stadiometers and Class III Seca scales. For interviews completed over the telephone, child height and weight were measured and reported by parents (if the family did not own a body weight scale, parents were asked to take their child to their local GP surgery or pharmacy prior to the telephone interview). Heights and weights were used to calculate body mass index standard deviation scores (BMI-SDS), adjusted for age and sex based on British 1990 growth reference data^33,34^.

#### Covariates

Parents reported their child’s age (years), sex, and ethnicity based on the 2021 list of ethnic groups used in the census survey for England and Wales. Child ethnicity was categorised as ‘White’, ‘Black, Black British, Caribbean, or African’, ‘Mixed or multiple ethnic groups’, ‘Asian or Asian British’, or ‘other ethnic group’.

### Statistical analysis

Analyses were conducted using R (version 4.1.1; R Foundation in Statistical Computing). Missing data on covariates, exposures (family-level SEP) and outcomes were imputed using Multivariate Imputation by Chained Equations (*mice)* package with a maximum of 50 iterations to create 20 imputed datasets^35^. All variables included in this analysis were used as predictors of imputed estimates. Pooled results from the imputed datasets are reported.

A series of Generalised Linear Models were used to examine cross-sectional associations between: (i) socioeconomic position and neighbourhood food environment as independent variables (IVs), and each family food-related outcome variable (dependent variables, DVs) in separate models (Model 1); and (ii) the interaction between socioeconomic position and neighbourhood food environment (IVs) and each DV in separate models (Model 2). Child-BMI was also examined as a DV in both models, to ensure SEP was associated with child adiposity, as expected, and explore associations with the food environment. Each model adjusted for child age, sex and ethnicity, and clustering of children at the school-level, using the *survey*^36^ package in R. Alpha was set at .05. All results are presented in full, with standard errors or 95% confidence intervals and p-values. Standardised betas are reported to provide comparable effect sizes for all analyses. Standardised betas ≥0.10 (and p<.05) were considered meaningful for family food-related variables, BMI and socioeconomic position variables.

#### Stakeholder and public involvement and engagement

The study’s Public Involvement and Engagement (PIE) group, the Caregiver Advisory Panel (CGAP; https://blogs.city.ac.uk/familyfoodexperiencestudy/cgap/), included six parents/caregivers who collaborated as research partners. The CGAP were involved through eight two-hour online group meetings, each of which was guided by two or three ‘real-time’ project sensitive questions suggested by the core FFES-L team. One additional meeting was held to get feedback from the CGAP on the interpretation of preliminary results. The meetings were facilitated by the FFES-L PPI-lead (CS) and either DB or SP (full names in Acknowledgments), or CV and AK for one final PPI input meeting. As required, another member of the wider FFES-L project team or a representative from the communications and marketing company we used attended too. The CGAP contributed to the design of recruitment and data collection methods, and interpretation of results. Based on feedback from CGAP members of their experiences of how their neighbourhood food environments affect their health and the broader neighbourhood food environment literature^24^, the effect size cut-off for neighbourhood food environment results was set at a less stringent 0.05 and p<0.05. PIE in this paper is reported in line with the 5-item GRIPP2 (short form) international reporting guidelines^37^ plus three additional items recommended by subsequent research^38^. The FFES-L core research team also conducted a series of one-on-one interviews with stakeholders (policymakers, strategic members of Local Authorities and health charities) prior to initiation of the study to co-develop overarching objectives.

This study was conducted according to the guidelines laid down in the Declaration of Helsinki and all procedures involving research study participants received ethical approval from City, University of London, School of Health & Psychological Sciences Ethics Committee (ID 2021-0367 & ID2021-1377).

## RESULTS

### Study population

The sample characteristics (n=728) overall and by borough are shown in **Table 1**. Most respondents were mothers or female caregivers (92.9%). Children were 4-11 years old (mean=7.29 years; SD=2.05), and approximately half were female (51.5%). The sample were ethnically diverse, including White (46.8%), Black, Black British, Caribbean, or African (22.8%), Mixed or multiple ethnic groups (18.7%), Asian or Asian British (8.0%), and ‘other’ ethnic group (2.7%).

**Table 1.** Characteristics of FFES-L, by London borough and for the overall sample (n=728)

| Characteristics | Borough |  |  |  | Full sample<br>Mean (SD) or N (%)<br>(n=728) |
| --- | --- | --- | --- | --- | --- |
|  | Haringey<br>Mean (SD) or N (%)<br>(n=180) | Croydon<br>Mean (SD) or N (%)<br>(n=208) | Greenwich<br>Mean (SD) or N (%)<br>(n=147) | Southwark<br>Mean (SD) or N (%)<br>(n=193) |  |
| Sex of parent, No. (% female) | 170 (94.4) | 192 (92.3) | 134 (91.2) | 180 (93.3) | 676 (92.9) |
| Age of parent | 40.17 (6.93) | 39.14 (5.86) | 40.18 (5.95) | 40.56 (6.96) | 39.90 (6.57) |
| **Ethnicity, No. (%)** |  |  |  |  |  |
| White | 80 (44.4) | 96 (46.2) | 80 (54.4) | 85 (44.0) | 341 (46.8) |
| Mixed or multiple ethnic groups | 31 (17.2) | 42 (20.2) | 31 (21.1) | 32 (16.6) | 136 (18.7) |
| Asian or Asian British | 7 (3.9) | 23 (11.1) | 13 (8.8) | 15 (7.8) | 58 (8.0) |
| Black, Black British, Caribbean, or African | 54 (30.0) | 43 (20.7) | 20 (13.6) | 56 (29.0) | 173 (23.8) |
| Other ethnic group | 8 (4.4) | 4 (1.9) | 3 (2.0) | 5 (2.6) | 20 (2.7) |
| Relationship to child, No. (%) birth parent | 176 (97.8) | 202 (97.1) | 143 (97.3) | 191 (99.0) | 712 (97.8) |
| **Maternal educational qualifications<sup>1</sup>** |  |  |  |  |  |
| Low | 39 (21.8) | 28 (13.5) | 20 (13.6) | 28 (14.5) | 115 (15.8) |
| Middle | 63 (35.2) | 56 (26.9) | 41 (27.9) | 56 (29.0) | 216 (29.7) |
| High | 77 (43.0) | 124 (59.6) | 86 (58.5) | 109 (56.5) | 396 (54.5) |
| **NS-SEC<sup>2</sup>** |  |  |  |  |  |
| Low | 110 (61.1) | 63 (30.3) | 43 (29.3) | 66 (34.2) | 282 (38.7) |
| Middle | 12 (6.7) | 22 (10.6) | 18 (12.2) | 36 (18.7) | 88 (12.1) |
| High | 58 (32.2) | 123 (59.1) | 86 (58.5) | 91 (47.2) | 358 (49.2) |
| **Housing tenure<sup>3</sup>** |  |  |  |  |  |
| Own without mortgage | 7 (3.9) | 7 (3.4) | 9 (6.1) | 10 (5.2) | 33 (4.5) |
| Own with mortgage | 34 (18.9) | 106 (51.0) | 61 (41.5) | 60 (31.1) | 261 (35.9) |
| Rent privately | 74 (41.1) | 48 (23.1) | 33 (22.4) | 37 (19.2) | 192 (26.4) |
| Rent from local authority | 65 (36.1) | 47 (22.6) | 43 (29.9) | 86 (44.6) | 242 (33.2) |
| **Index of Multiple Deprivation<sup>4</sup>** |  |  |  |  |  |
| 1 – most deprived | 99 (55.0) | 54 (26.1) | 36 (24.5) | 56 (29.0) | 245 (33.7) |
| 2 | 46 (25.6) | 44 (21.3) | 52 (35.4) | 76 (39.4) | 218 (30.0) |
| 3 | 19 (10.6) | 67 (32.4) | 29 (19.7) | 43 (22.3) | 158 (21.7) |
| 4 | 11 (6.1) | 22 (10.6) | 28 (19.0) | 14 (7.3) | 75 (10.3) |
| 5 – least deprived | 5 (2.8) | 20 (9.7) | 2 (1.4) | 4 (2.1) | 31 (4.3) |
| **Annual household income** |  |  |  |  |  |
| Low (<£30,000 per year) | 97 (53.9) | 59 (28.4) | 49 (33.3) | 97 (50.3) | 302 (41.5) |
| Medium (£30,000-67,500 per year) | 35 (19.4) | 60 (28.8) | 38 (25.9) | 44 (22.8) | 177 (24.3) |
| High (>£67,500 per year) | 48 (26.7) | 89 (42.8) | 60 (40.8) | 52 (26.9) | 249 (34.2) |
| Number of bedrooms | 2.42 (1.02) | 2.94 (0.99) | 2.84 (0.86) | 2.54 (1.02) | 2.69 (1.00) |
| Number of cars | 0.54 (0.66) | 1.06 (0.72) | 0.80 (0.66) | 0.59 (0.60) | 0.76 (0.70) |
| Living w/partner (as couple) in house, N (%) | 92 (51.1) | 140 (67.3) | 102 (69.4) | 110 (57.0) | 444 (61.0) |

| Characteristics | Borough |  |  |  | Full sample |
| --- | --- | --- | --- | --- | --- |
|  | Haringey<br>Mean (SD) or N<br>(%) (n=180) | Croydon<br>Mean (SD) or N (%)<br>(n=208) | Greenwich<br>Mean (SD) or N (%)<br>(n=147) | Southwark<br>Mean (SD) or N (%)<br>(n=193) |  |
| Overall SEP composite<sup>5</sup> | 3.32 (1.61) | 4.33 (1.61) | 4.35 (1.70) | 3.88 (1.66) | 3.96 (1.69) |
| Sex of child, No. (% female) | 100 (55.6) | 102 (49.0) | 75 (51.0) | 98 (50.8) | 375 (51.5) |
| Age of child, ranged from 4-11 years | 7.62 (1.98) | 7.20 (2.14) | 7.03 (2.02) | 7.26 (2.02) | 7.29 (2.05) |
| BMI-SDS<sup>6</sup> | 0.52 (1.73) | 0.06 (1.61) | 0.39 (1.28) | 0.59 (1.38) | 0.41 (1.54) |
| **Mode of interview delivery** |  |  |  |  |  |
| *Face to face* | 39 (21.7) | 33 (15.9) | 117 (79.6) | 35 (18.1) | 224 (30.8) |
| *Telephone* | 141 (78.3) | 175 (84.1) | 30 (20.4) | 158 (81.9) | 504 (69.2) |
| *Interview length (in minutes)* | 78.51 (49.01) | 58.22 (33.36) | 68.16 (30.40) | 64.34 (27.38) | 67.01 (39.09) |

<sup>1</sup> Education level was categorised as: low (no qualifications or high school education e.g. CSE, GCSE, O level), intermediate (vocational qualification or advanced high school education), and high (University-level education). Data were available for 727 participants (n=1 case missing).
<sup>2</sup> Classified based on the Office for National Statistics Socioeconomic Classification (NS-SEC) and grouped into high (higher and lower managerial and professional occupations), middle (intermediate occupations, small employers and own account workers) and low (lower supervisory and technical occupations, (semi)routinised occupations, never worked and long-term unemployed).
<sup>3</sup> Annual household income information was available for 653 families (n=76 missing).
<sup>4</sup> Index of multiple deprivation could be calculated for 727 families (n=1 missing).
<sup>5</sup> SEP-composite score was based on a weighted score using seven indicators of socioeconomic position. These included maternal educational qualifications, household NS-SEC score, home ownership status, annual household income, household composition e.g. bedrooms and cars. <sup>21</sup>. n=78 could not be classified due to missing data on at least one variable within the composite score.
<sup>6</sup> Standard deviation scores (SDS) for child height, weight, and body mass index (BMI) were calculated using the UK1990 British growth reference data, adjusting for child age and sex. Data were available for 635 (86.3%) of participants (n=101 cases missing).

### Deprivation (Socioeconomic Position, SEP composite measure) of the study population

The sample were socioeconomically diverse, with 38.7% living in the most deprived areas based on IMD quintiles, and 28.3% reporting low or very low food security. Representativeness of the overall study sample compared to wider Local Authority-level data on ethnicity, child weight status and IMD are shown in **Table S1**. Most participants (69.2%) completed the interview via telephone. The average length of interview was 67 minutes (SD = 39 minutes).

### Neighbourhood food environment characteristics

The median neighbourhood food environment score was −59.0, ranging from −1349.0 to 142.0 (IQR: −119.0, −15.5). An example of the food outlet exposure for a child with a score of –59.0 is shown in **Figure S2.** Almost all children (87.3%, n=630) had a negative food environment score, indicating greater exposure to food outlets selling predominantly less healthy foods. 22

#### Overall Findings

Fully adjusted models showing the results of associations between deprivation and the neighbourhood food environment with home food environments, parental feeding practices, and child eating behaviours, food preferences and BMI, and the interactive effects between SEP and neighbourhood food environment on these dependent variables, are presented in **Table 2**.

**Table 2.** Generalised Linear Models examining associations between socioeconomic position, the neighbourhood food environment (NFE), and their interaction, with the home food environment, parental feeding practices, and child eating behaviours, food preferences and BMI-SDS (n=728).

| Outcomes | SEP composite |  | Neighbourhood food environment (NFE) |  | Interaction: SEP * NFE |  |
| --- | --- | --- | --- | --- | --- | --- |
| | $\beta$ (95% CI) | p-value | $\beta$ (95% CI) | p-value | $\beta$ (95% CI) | p-value |
| **Home food environment** | **-0.19 (-0.09, -0.28)** | **<0.001** | **-0.07 (-0.02, -0.13)** | **<0.01** | 0.01 (-0.05, 0.07) | 0.713 |
| **Parental feeding practices** |  |  |  |  |  |  |
| Emotional feeding | **-0.10 (-0.20, -0.01)** | **0.031** | 0.01 (-0.08, 0.10) | 0.876 | 0.08 (-0.02, 0.17) | 0.107 |
| Instrumental feeding | **-0.11 (-0.04, -0.18)** | **0.003** | 0.01 (-0.04, 0.06) | 0.722 | 0.07 (-0.03, 0.16) | 0.163 |
| Encouragement to eat | -0.03 (-0.11, 0.05) | 0.455 | 0.02 (-0.06, 0.11) | 0.586 | -0.03 (-0.13, 0.06) | 0.472 |
| Modelling | **0.10 (0.00, 0.20)** | **0.044** | 0.02 (-0.10, 0.14) | 0.735 | 0.02 (-0.09, 0.14) | 0.666 |
| Monitoring | -0.04 (-0.14, 0.05) | 0.348 | 0.00 (-0.08, 0.09) | 0.943 | 0.01 (-0.08, 0.10) | 0.800 |
| Covert restriction | 0.01 (-0.09, 0.11) | 0.785 | -0.01 (-0.11, 0.09) | 0.816 | -0.02 (-0.13, 0.09) | 0.748 |
| Restriction | 0.10 (-0.01, 0.20) | 0.070 | 0.04 (-0.05, 0.14) | 0.373 | -0.01 (-0.12, 0.10) | 0.838 |
| Parent Control | **0.17 (0.09, 0.24)** | **0.001** | -0.01 (-0.09, 0.07) | 0.725 | 0.05 (-0.06, 0.17) | 0.325 |
| Pressure to eat | **-0.14 (-0.21, -0.07)** | **0.001** | 0.00 (-0.09, 0.09) | 0.991 | 0.05 (-0.05, 0.15) | 0.264 |
| Mealtime structure | -0.00 (-0.08, 0.08) | 0.977 | -0.01 (-0.07, 0.05) | 0.809 | 0.06 (-0.03, 0.14) | 0.159 |
| **Child Eating behaviours** |  |  |  |  |  |  |
| Food responsiveness | **-0.12 (-0.04, -0.20)** | **0.002** | 0.03 (-0.02, 0.07) | 0.212 | -0.05 (-0.12, 0.02) | 0.176 |
| Enjoyment of food | 0.01 (-0.07, 0.09) | 0.776 | 0.07 (-0.01, 0.14) | 0.070 | -0.06 (-0.13, 0.01) | 0.092 |
| Emotional overeating | **-0.11 (-0.05, -0.17)** | **<0.001** | -0.02 (-0.08, 0.04) | 0.550 | 0.03 (-0.04, 0.09) | 0.384 |
| Desire to drink | **-0.26 (-0.33, -0.17)** | **<0.001** | -0.03 (-0.09, 0.03) | 0.355 | -0.03 (-0.11, 0.05) | 0.451 |
| Satiety responsiveness | -0.00 (-0.09, 0.09) | 0.962 | -0.06 (-0.16, 0.03) | 0.178 | 0.05 (-0.08, 0.18) | 0.395 |
| Slowness in eating | -0.06 (-0.15, 0.02) | 0.135 | -0.04 (-0.14, 0.05) | 0.346 | 0.04 (-0.09, 0.16) | 0.496 |
| Emotional undereating | -0.05 (-0.14, 0.03) | 0.198 | -0.02 (-0.09, 0.05) | 0.582 | 0.03 (-0.08, 0.13) | 0.584 |
| Food fussiness | 0.01 (-0.06, 0.09) | 0.778 | -0.06 (-0.16, 0.04) | 0.272 | 0.01 (-0.12, 0.14) | 0.867 |
| **Child food preferences** |  |  |  |  |  |  |
| Fruit | 0.03 (-0.04, 0.11) | 0.393 | 0.08 (-0.00, 0.16) | 0.056 | 0.02 (-0.06, 0.09) | 0.681 |
| Vegetables | 0.03 (-0.05, 0.11) | 0.529 | 0.05 (-0.06, 0.15) | 0.375 | -0.05 (-0.17, 0.07) | 0.411 |
| Protein (meat/fish) | 0.05 (-0.04, 0.14) | 0.298 | 0.02 (-0.04, 0.09) | 0.507 | 0.02 (-0.05, 0.09) | 0.575 |
| Starch | **0.10 (0.02, 0.17)** | **0.015** | 0.08 (-0.05, 0.20) | 0.232 | -0.01 (-0.11, 0.10) | 0.873 |
| Dairy | 0.03 (-0.04, 0.11) | 0.379 | 0.04 (-0.05, 0.13) | 0.373 | 0.00 (-0.09, 0.09) | 0.990 |
| Snacks | **0.15 (0.07, 0.23)** | **0.001** | 0.06 (-0.01, 0.13) | 0.110 | 0.01 (-0.06, 0.09) | 0.754 |
| **Child BMI-SDS** | **-0.23 (-0.06, -0.39)** | **0.007** | -0.09 (-0.34, 0.17) | 0.443 | -0.03 (-0.17, 0.13) | 0.725 |

\*Models are adjusted for child age, child sex, and child ethnicity and for clustering at the school-level. Standardised betas with an effect size $\geq 0.10$ , that meet the alpha level $<0.05$ are shown in bold.

#### Overall findings – Socioeconomic position

Greater deprivation was associated with living in a more obesogenic home food environment (β= −0.19; 95% CI = −0.09, −0.28; p<.001). When examining parental feeding practices, greater deprivation was associated with greater use of emotional feeding (β= −0.10; −0.01, −0.20; p=.031) and instrumental feeding (β= −0.11; −0.04, −0.18; p=.003), greater pressure to eat (β= −0.14; −0.07, −0.21; p=0.001), less modelling of healthy eating (β= 0.10; 0.00, 0.20; p=.044), and less control/structure over meals and snack times (β=0.17; 0.09, 0.24; p<.001). For child eating behaviours, greater deprivation was associated with higher responsiveness to food cues (β= −0.12; −0.04, −0.20; p=0.002), higher emotional over-eating (β= −0.11; −0.05, −0.17, p<0.001), and higher desire to drink (β= −0.26; −0.17, −0.33; p<0.001). It was also significantly associated with lower preference for snack foods and starchy foods (β= 0.15; 0.07, 0.23; p=0.001 and β=0.10; 0.02, 0.17; p=0.015, respectively). As expected, greater deprivation (β= −0.23; −0.06, −0.39; p=.007) was associated with higher child BMI-SDS. All associations were small or small-to-moderate in magnitude.

**Fig 1.**
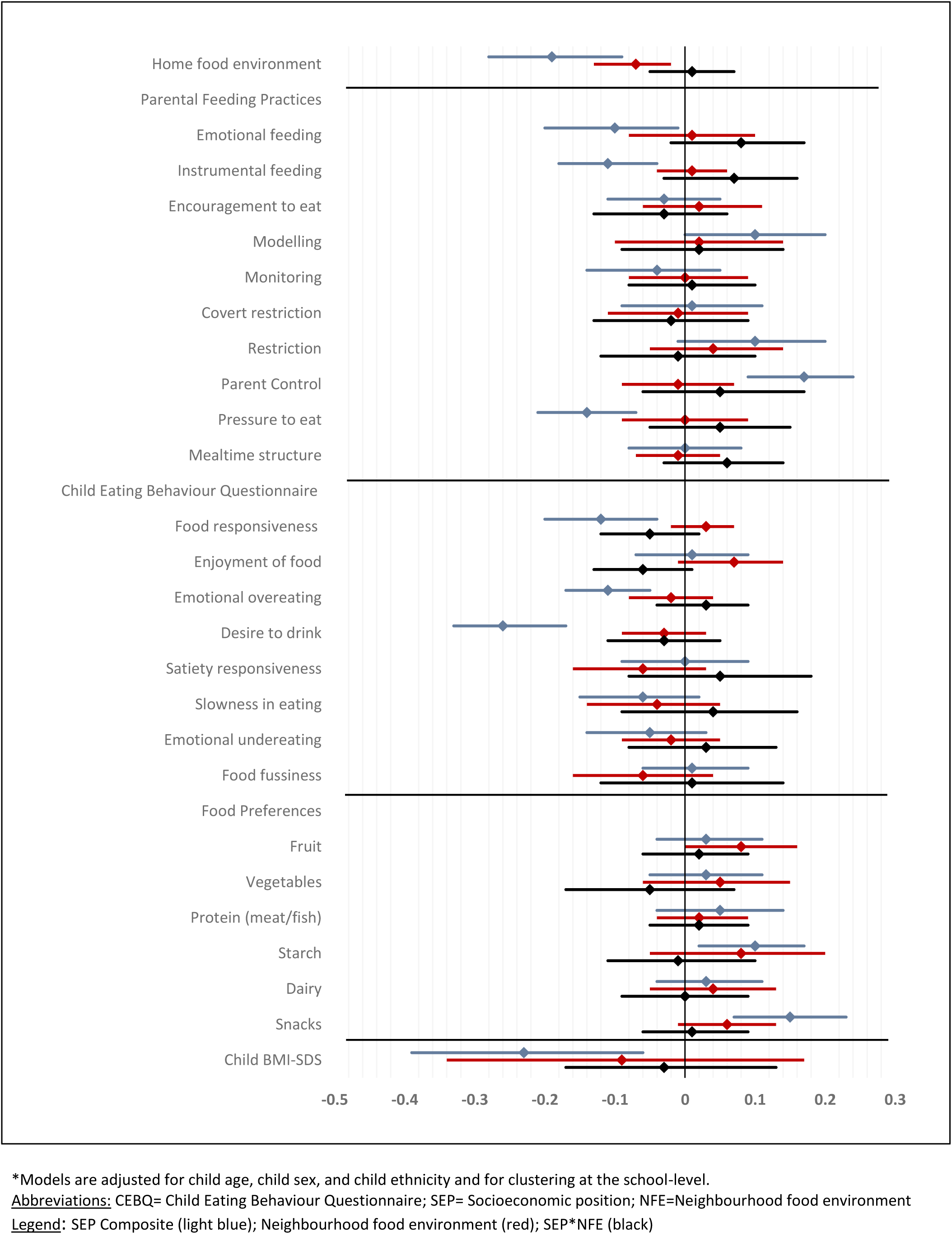
Associations between socioeconomic position (blue), the neighbourhood food environment (red)), and their interaction (green), with the home food environment, parental feeding practices, child eating behaviours, child food preferences and child BMI-SDS (n=728)

#### Overall findings – Neighbourhood food environment

Exposure to a less healthy neighbourhood food environment was significantly associated with living in a more obesogenic home food environment, (β= −0.07; −0.02, −0.13; p<0.01). No associations were observed between the neighbourhood food environment and any parental or child food-related variables including child BMI-SDS. No significant interactions were observed between SEP and the neighbourhood food environment with the parental or child food-related variables including child BMI-SDS (see **Table 2**).

#### Evidence of Stakeholder and Public involvement and engagement (PIE)

PIE shaped this study via discussions on key project-sensitive topics. In the first CGAP meeting, participants provided new ideas to increase engagement by schools and families, such as forging stronger partnerships with LA and tailoring communication to diverse demographic groups. CGAP members also suggested changes to participant recruitment and information materials to simplify them, build trust, and widen access (e.g. highlighting that some participants would not be able to access study materials on a computer, such that all materials would need to be viewable on a mobile phone). CGAP members also provided ideas for participation incentives, including monetary and non-monetary rewards, such as gift cards, vegetable boxes and public recognition (by means of a letter for participating children). In a separate meeting, discussions expanded to include recruitment suggestions such as using parent champions and developing a short, fun, and bright recruitment video. Additionally, a newly developed evidence-based ‘Top Tips for Healthy Eating’ flyer was developed in response to requests from CGAP members as many parents find it difficult to identify reliable healthy eating information. This flyer also served as a thank-you to those who completed the interview.

## DISCUSSION

This study contextualised, for the first-time, a comprehensive range of family food norms and children’s obesity-related behaviours, according to deprivation level and the proximal neighbourhood food environment. The London-based sample showed that greater neighbourhood deprivation was related to living in a more obesogenic neighbourhood and home food environment, and more frequent use of non-nutritive and less structured caregiver feeding practices. Living in more deprived neighbourhoods also expressed more obesogenic eating behaviours, showed differences in some food preferences, and had a higher BMI. Less healthy neighbourhood food environments were associated with a more obesogenic home food environment, but no other effects for neighbourhood food environment were observed. These findings indicate that some aspects of family food culture and the home food environment vary by level of family deprivation and neighbourhood food environment exposures, highlighting the need for LAs to take the context of deprivation into account when developing interventions to address inequalities in childhood obesity.

### Findings in Context

Family-level deprivation is one of the most important determinants of childhood obesity in the UK ^1^. The results of this study confirm national trends showing that primary school-aged children from more disadvantaged backgrounds had a higher BMI than children from more affluent backgrounds. Our findings provide important new insights into how the context of deprivation creates barriers to families engaging with, and benefitting from, local interventions aimed at reducing inequalities in childhood obesity. Specifically, we identified several modifiable family food-related domains and parent-child behaviours that varied with deprivation. The findings also emphasized the need for targeted interventions in the future that address both the broader food environment and household-specific food practices used by families living in household deprivation. The results also re-emphasize that systems-thinking approach is essential to tackling these disparities, as it considers the complex interplay between individual behaviours, family dynamics, community resources, and policy-level influences, ensuring that interventions are both comprehensive and sustainable.

In this study, greater deprivation was associated with the expression of some child eating behaviours that have been robustly associated with higher risk of obesity^20^. Namely: heightened responsiveness to food cues (’food responsiveness’); a greater tendency to want to eat in response to negative emotion (’emotional overeating’); and a stronger ‘desire to drink‘. These findings align with previous prospective research, which found that toddlers from more disadvantaged backgrounds had greater increases in food responsiveness and emotional overeating from toddlerhood to early^21^. However, deprivation was also associated with lower preferences for snack and starchy foods among children. These findings may, in part, be explained by the neighbourhood environment findings, which indicate that children living in areas had greater exposure to less healthy food outlets and less healthy food environments at home. Children’s eating styles may be reflecting their food environments, rather than their food preferences. Children in more deprived areas may show a lower reported preference for snacks and starchy foods because these items are widely available and routinely consumed, making them less of a “treat” compared to how they are perceived by more affluent children, who may have them less frequently but are still influenced by marketing ^39^. Additionally, parental reporting biases could play a role, as more affluent parents may perceive their child’s preference for these foods as stronger in contrast to healthier options that are more accessible to them, whereas lower-income parents may view such preferences as less pronounced since healthier alternatives are often not a feasible option. This situation may therefore pose barriers to these children receiving benefits from healthy and drinking eating interventions in the London area.

Previous studies have observed that children from more disadvantaged backgrounds are more likely to live in higher risk ‘obesogenic environments’, with more access to unhealthy foods and greater exposure to environmental cues to eat (such as higher density of unhealthy food outlets, food advertisements, etc.)^40^. This was also observed in our study, with greater deprivation being significantly associated with both a more obesogenic home food environment and greater exposure to less healthy neighbourhood food environments. Greater exposure to these environments may play a role in nurturing the obesogenic eating behaviours observed in this study and may contribute to the socioeconomic differences in these traits. However, our findings suggest that the neighbourhood food environment was not directly associated with child eating behaviours, indicating that the relationship may be more complex and multifaceted, potentially involving other environmental or individual factors. These findings were also seen in a study of the neighbourhood food environment of ethnic minority groups in Amsterdam, which found was less healthy and diverse than that of Dutch-origin participants^41^. The authors of the HELIUS study (n=4,728) reported that no evidence was found that it contributed to ethnic differences in diet quality, which suggests individuals and families interact with their food environments in different ways not entirely captured by indices of environmental exposure alone. This is because it is only one dimension of the food environment which does not consider price or marketing activities such as placement, promotion or advertising^42^. There is evidence that the neighbourhood food environment is a lower influence on diet than consumer food environment ^43^. Additionally, the vast majority of children in these four boroughs of London lived in an ‘unhealthy’ neighbourhood food environment (NFE score <0). This lack of variability in the sample, is likely to have reduced the ability to detect statistically significant effects. It is important to replicate these findings in a sample with greater variation in neighbourhood food environment quality and examine relationships in a range of rural and urban contexts.

Parents and caregivers from more deprived backgrounds participating in this study reported more frequent use of some parental feeding practices that have previously been associated with higher BMI in childhood^44^. These approaches included greater use of non-nutritive feeding practices, such as emotional feeding (e.g., using food to soothe negative emotions) and instrumental feeding (e.g., using food to reward or punish behaviour), as well as less structure/control over meals and snack times. Greater household deprivation was also associated with a more pressuring feeding style. Previous research has also that parents from more disadvantaged backgrounds report using food as a reward or comforter for their child more frequently than more affluent parents^45–47^. These differing parental approaches may arise from their financial circumstances. Mothers living on lower incomes have previously described the importance of not wasting food (pressuring to eat) and the joy that eating affordable foods, like takeaways and confectionary, brings their family when other rewards such as toys or day trips are unaffordable for them^39,48^. These findings highlight that parental feeding practices are not simply about nutrition but are frequently driven by the contexts of deprivation which leave families living in more difficult socioeconomic circumstances with more limited choices.

The neighbourhood food environment showed no relationship with child weight status or any of the family food-related outcomes assessed in this study (e.g. parental feeding practices, child eating behaviours, child food preferences). Previous research on the impact of the neighbourhood food environment on child and parent behaviours are mixed^8,9,49^. This inconsistency may reflect the interplay between environmental exposure and individual characteristics (such as differences in eating styles), which ‘muddies the water’ when examining population averages^50,51^. Pathway modelling of a range of environmental and individual-level characteristics in future research may help to identify the relative importance of environmental, social and individual determinants of childhood obesity and help to pinpoints key target areas in future interventions^52^.

### Implications

The overall aim of the FFES-L study was to understand how the context of deprivation prevents LA interventions and policies from effectively reducing social disparities in childhood obesity. The socioeconomic patterning at individual, household and neighbourhood levels identified in this study are likely to have an accumulative contribution to the well-established socioeconomic gradient in childhood obesity. It is unlikely that LA interventions targeting childhood obesity have managed to specifically address all levels of drivers and therefore have been ineffective. For example, providing families with financial or practical support to afford healthy foods is important but providing other forms of leisure activities and social support may be important for families living in greater deprivation to support them in minimising the use of food for non-nutritive purposes (e.g. to soothe negative emotions). These circumstances not only shape children’s food preferences but also reinforce food practices and parental feeding styles, as parents in deprived settings may be more likely to use less healthy foods to meet both nutritional and emotional needs, particularly when other ways of expressing care and providing treats are financially out of reach (Parish et al, *Under Review*) ^39^. The association between neighbourhood and family food environments observed in this study highlights how the contexts in which children experiencing greater deprivation live exacerbate responsiveness to unhealthy environmental food cues. It is crucial for LAs to take account of the neighbourhood food environment (particularly where children live, commute and play) in interventions to enable them to most effectively reduce inequalities in childhood obesity. A modelling study provides evidence that takeaway management zones around schools reduce the number of hot food takeaways, potentially decreasing exposure and leading to significant reductions in adult obesity prevalence and related health burdens ^53^. This approach is feasible for wider adoption. Future research should explore the feasibility and impact of ‘school superzones’ (a 400 m zone around a school to reduce harmful exposures such as takeaway outlets) and strategies to improve the healthiness of small and medium-sized enterprises (e.g. corner shops), which are largely exempt from current UK government food policies. Evaluations of these initiatives will help determine their effectiveness in improving neighbourhood food environments.

LAs often rely on using existing interventions (i.e., take things ‘off the shelf’) and applying them in their communities with very few resources^54^. However, they also recognise the need to adapt and augment interventions to meet the needs of the communities they serve^55^. Our findings suggest three important recommendations for LAs to consider when adapting existing healthy eating interventions which aim to reduce inequalities in childhood obesity. Firstly, drinking behaviours are as important as eating behaviours in programs aimed at improving child nutrition and weight, and are socio-economically patterned. Children from more disadvantaged backgrounds expressed both a greater compulsion to eat in response to external food cues, and a stronger desire to consume drinks. Both behaviours can contribute to excess intake of low nutrient-dense items (in 2021 45% of 11–18-year-olds in England consume soft drinks) ^56^; and both tendencies pose barriers to the implementation of healthy eating and drinking initiatives. Making healthy food affordable and accessible to families is important, but it may not be enough if children are still surrounded by cues (e.g. food advertising) to consume food and drinks high in fat, salt or sugar (HFSS), which are difficult to resist and cheaper ^57,58^. In the context of childhood obesity, effective approaches include water-only schools and sugar-sweetened beverage drinks levys^58,59^. Secondly to tailor interventions to ‘the world view’ of the target community^60^ in recognition that many families use food not just for nutrition, but also to show care, soothe emotions or connect with their children in affordable ways. Interventions are more likely to be effective if they are realistic, feasible and compatible with the complex lives of families. Thirdly, to address the link between the neighbourhood food environment and the home family food environment by taking action to improve the healthfulness of the wider environment. Families experiencing greater deprivation are often more dependent on their local neighbourhood food outlets than more affluent families ^20^. It is therefore important that interventions to address inequalities in childhood obesity making healthier choices more accessible, more affordable, and easier for all families to achieve. Action in this area is more important than ever, given the rise in the cost of living and rates of food insecurity in England^61^. This final recommendation recognises that family food practices that influence childhood diet and obesity are shaped by broader environmental and social drivers which require ‘upstream’ policies to enable any ‘downstream’ policies targeting families and individuals to be effective.

#### Reflections on public involvement and engagement

PIE in this study provided the researchers with practical suggestions for recruitment strategies that may otherwise have been overlooked, such as offering a mixture of monetary incentives to parents and non-monetary rewards to children (e.g. letters/certificates for participating), as well as mobilising established parent champions in schools and communities. We believe adoption of these suggestions, provided by the CGAP, increased recruitment. Co-producing the aims of the analyses with stakeholders in local government (LAs/Boroughs), public contributors, and in-depth qualitative interviews with families meant the analyses addressed the most pertinent questions to organisations who are most committed to addressing childhood obesity, and to communities that would benefit from the interventions.

#### Strengths and Limitations

Study strengths include the relatively large, ethnically and socioeconomically diverse sample, and use of a comprehensive battery of validated psychometric measures. Importantly, PIE was involved throughout the entire study which helped to shape the study materials, methods and interpretation of the results to ensure they were appropriate, and representative of families’ lived experiences. Nonetheless, there were several limitations. Firstly, family food-related outcomes were parent-reported, which makes them subjective and susceptible to desirability bias. Secondly, the study was cross-sectional, which means it is not possible to draw conclusions about the direction of observed relationships and causality cannot be established between the exposures and outcomes. While it is not plausible that family food-related factors can directly cause socioeconomic position, other unmeasured confounding factors could be causally involved (such as parental obesity). Thirdly, although the sample was ethnically and socioeconomically diverse, the population was limited to families living in four London boroughs, therefore the results cannot be replicated to other areas of the UK (e.g. rural areas in other parts of the country) or internationally. Furthermore, London is a very densely populated area, with pockets of people living with greater deprivation close to areas with people not experiencing poverty, and the neighbourhood food environment exposure may perform differently in less densely and less transient populated areas within the United Kingdom. Further research is needed in a diverse sample of families from across the UK, with greater variation in the neighbourhood food environment and potentially examining pathways between multiple environmental and individual levels factors. Additionally future research may wish to examine child and parental neighbourhood food environment exposures to understand the direct and indirect effects on outcomes such as the home food environment and parental feeding practices.

#### Conclusion

This study highlights that family-level deprivation, rather than neighbourhood food environments, is closely linked to more ‘obesogenic’ parental food practices, child food behaviours, preferences, and BMI, suggesting that policies focused on ‘out of home’ interventions may have small but adjunctive impact amidst rising deprivation to consider alongside broader strategies. Policymakers might consider addressing welfare policies—such as universal credit, benefit caps, and the two-child limit— as these forms of support could mitigate some barriers to engaging with local healthy eating interventions. To effectively reduce social inequalities in childhood obesity, future research is needed to examine if the effectiveness of existing LA interventions can be improved, if they are adapted and augmented to take these factors into account.

## Data Availability

All data produced in the present study are available upon reasonable request to the authors

## Data Availability

Data are available in the UK Data Service and available on application to the UK Data Service (https://ukdataservice.ac.uk/).

## Abbreviations

## Acknowledgements

We thank all families who participated in the Family Food Experience Study – London, NatCen who undertook data collection, and the study’s Public Involvement and Engagement (PIE) group, the Caregiver Advisory Panel (CGAP; https://blogs.city.ac.uk/familyfoodexperiencestudy/cgap/), included six core members who were collaboratively involved as research partners. We would also like to thank Professor Carolyn Summerbell (CS), Dr Daisy Bradbury (DB) and Dr Sabine Parrish (SP) who facilitated the meetings with the CGAP and were instrumental in gaining this important PIE input. We also acknowledge the support and input from the wider core management group of the FFES-L study including Professor Tessa Pollard, Adrian White, Dr Anna Isaacs, Dr Mark Spires, Dr Kyriaki Giorgakoudi, Dr Natalie Savona, Dr Charitini Stavropoulou and Professor Corinna Hawkes. This study was supported by the National Institute of Health Research, grant number NIHR129771.

## REFERENCES

1. NHS Digital. National Child Measurement Programme, England, 2023/24 School Year - NHS England Digital. https://digital.nhs.uk/data-and-information/publications/statistical/national-child-measurement-programme/2023-24-school-year (2024).

2. House of Lords. Recipe for Health: A Plan to Fix Our Broken Food System. (2024).

3. Joseph Rowntree Foundation. UK Poverty 2024: The essential guide to understanding poverty in the UK. https://www.jrf.org.uk/uk-poverty-2024-the-essential-guide-to-understanding-poverty-in-the-uk (2024).

4. NHS Digital. National Child Measurement Programme, England, 2022/23 School Year. https://digital.nhs.uk/data-and-information/publications/statistical/national-child-measurement-programme/2022-23-school-year/deprivation-based-on-postcode-of-the-child (2023).

5. Taylor-Robinson, D. C., Lai, E. T., Whitehead, M. & Barr, B. Child health unravelling in UK. BMJ 364, (2019).

6. Public Health England. Whole systems approach to obesity: A guide to support local approaches. (2019).

7. Taheem, R., Woods-Townsend, K., Lawrence, W., Baird, J. & Godfrey, K. M. How do local authority plans to tackle obesity reflect systems thinking? Perspect Public Health 143, 324–336 (2023).

8. Engler-Stringer, R., Le, H., Gerrard, A. & Muhajarine, N. The community and consumer food environment and children’s diet: A systematic review. BMC Public Health 14, 1–15 (2014).

9. Black, C., Moon, G. & Baird, J. Dietary inequalities: What is the evidence for the effect of the neighbourhood food environment? Health Place 27, 229–242 (2014).

10. Atanasova, P. et al. The impact of the consumer and neighbourhood food environment on dietary intake and obesity-related outcomes: A systematic review of causal impact studies. Soc Sci Med 299, 114879 (2022).

11. Public Health England. Obesity and the environment - Density of fast food outlets at 31/12/2017. https://www.gov.uk/guidance/phe-data-and-analysis-tools#obesity-diet-and-physical-activity (2018) doi:10.1136/archdischild-2017-312981.

12. Maguire, E. R., Burgoine, T. & Monsivais, P. Area deprivation and the food environment over time: A repeated cross-sectional study on takeaway outlet density and supermarket presence in Norfolk, UK, 1990–2008. Health Place 33, 142–147 (2015).

13. Wijlaars, L. P. M. M., Johnson, L., Van Jaarsveld, C. H. M. & Wardle, J. Socioeconomic status and weight gain in early infancy. Int J Obes (Lond) 35, 963–970 (2011).

14. Kininmonth, A. R. et al. The Home Environment Interview and associations with energy balance behaviours and body weight in school-aged children – A feasibility, reliability, and validity study. International Journal of Behavioral Nutrition and Physical Activity (2022) doi:10.1186/s12966-021-01235-3.

15. Schrempft, S., Van Jaarsveld, C. H. M., Fisher, A. & Wardle, J. The obesogenic quality of the home environment: Associations with diet, physical activity, TV viewing, and BMI in preschool children. PLoS One 10, e0134490 (2015).

16. Adams, E. L., Caccavale, L. J., Larose, J. G., Raynor, H. A. & Bean, M. K. Home Food Environment Changes and Dietary Intake during an Adolescent Behavioral Weight Loss Intervention Differ by Food Security Status. Nutrients 14, (2022).

17. Costa, A. & Oliveira, A. Parental Feeding Practices and Children’s Eating Behaviours: An Overview of Their Complex Relationship. Healthcare 2023, Vol. 11, Page 400 11, 400 (2023).

18. Shloim, N., Edelson, L. R., Martin, N. & Hetherington, M. M. Parenting styles, feeding styles, feeding practices, and weight status in 4-12 year-old children: A systematic review of the literature. Front Psychol 6, 163321 (2015).

19. Mcphie, S., Skouteris, H., Daniels, L. & Jansen, E. Maternal correlates of maternal child feeding practices: A systematic review. Matern Child Nutr 10, 18–43 (2014).

20. Kininmonth, A. R. et al. The association between childhood adiposity and appetite assessed using the Child Eating Behavior Questionnaire and Baby Eating Behavior Questionnaire: A systematic review and meta-analysis. Obesity Reviews vol. 22 Preprint at 10.1111/obr.13169 (2021).

21. Kininmonth, A. R., Smith, A. D., Llewellyn, C. H. & Fildes, A. Socioeconomic status and changes in appetite from toddlerhood to early childhood. Appetite 146, 104517 (2020).

22. Scaglioni, S., Arrizza, C., Vecchi, F. & Tedeschi, S. Determinants of children’s eating behavior. Am J Clin Nutr 94, S2006–S2011 (2011).

23. Noble, S. et al. Indices of Deprivation 2019 Research Report. Ministry of Housing, Communities and Local Government 2–86 (2019).

24. Vogel, C. et al. The relationship between dietary quality and the local food environment differs according to level of educational attainment: A cross-sectional study. PLoS One 12, e0183700 (2017).

25. Moayyed, H., Kelly, B., Feng, X. & Flood, V. Evaluation of a ‘healthiness’ rating system for food outlet types in Australian residential communities. Nutrition & Dietetics 74, 29–35 (2017).

26. Birch, L. L. et al. Confirmatory factor analysis of the Child Feeding Questionnaire: A measure of parental attitudes, beliefs and practices about child feeding and obesity proneness. Appetite 36, 201– 210 (2001).

27. Wardle, J., Sanderson, S., Guthrie, C. A., Rapoport, L. & Plomin, R. Parental feeding style and the intergenerational transmission of obesity risk. Obes Res 10, 453–462 (2002).

28. Musher-Eizenman, D. & Holub, S. Comprehensive feeding practices questionnaire: Validation of a new measure of parental feeding practices. J Pediatr Psychol 32, 960–972 (2007).

29. Ogden, J., Reynolds, R. & Smith, A. Expanding the concept of parental control: A role for overt and covert control in children’s snacking behaviour? Appetite 47, 100–106 (2006).

30. Wardle, J., Guthrie, C. A., Sanderson, S. & Rapoport, L. Development of the Children’s Eating Behaviour Questionnaire. The Journal of Child Psychology and Psychiatry and Allied Disciplines 42, 963–970 (2001).

31. Carnell, S. & Wardle, J. Measuring behavioural susceptibility to obesity: Validation of the child eating behaviour questionnaire. Appetite 48, 104–113 (2007).

32. Fildes, A. et al. Nature and nurture in children’s food preferences. Am J Clin Nutr 99, 911–917 (2014).

33. Cole, T. J. Some Questions about How Growth Standards Are Used. Horm Res Paediatr 45, 18–23 (1996).

34. Freeman, J. V. et al. Cross sectional stature and weight reference curves for the UK, 1990. Arch Dis Child 73, 17–24 (1995).

35. Buuren, S. van. mice: Multivariate Imputation by Chained Equations in R. J Stat Softw 1–68 (2010) doi:10.18637/jss.v045.i03.

36. Lumley, T., Maintainer“, M. & Lumley”, T. Package ‘survey’ Title Analysis of Complex Survey Samples. (2021).

37. Staniszewska, S. et al. GRIPP2 reporting checklists: tools to improve reporting of patient and public involvement in research. BMJ 358, 3453 (2017).

38. Jones, J. et al. Reporting on patient and public involvement (PPI) in research publications: using the GRIPP2 checklists with lay co-researchers. Res Involv Engagem 7, 1–13 (2021).

39. The Food Foundation. From purse to plate: implications of the cost of living crisis on health. https://foodfoundation.org.uk/publication/purse-plate-implications-cost-living-crisis-health (2023).

40. Caldwell, A. E. & Sayer, R. D. Evolutionary considerations on social status, eating behavior, and obesity. Appetite 132, 238–248 (2019).

41. Poelman, M. P. et al. Does the neighbourhood food environment contribute to ethnic differences in diet quality? Results from the HELIUS study in Amsterdam, the Netherlands. Public Health Nutr 24, 5101–5112 (2021).

42. Glanz, K., Sallis, J. F., Saelens, B. E. & Frank, L. D. Healthy nutrition environments: concepts and measures. Am J Health Promot 19, 330–333 (2005).

43. Vogel, C. et al. Examination of how food environment and psychological factors interact in their relationship with dietary behaviours: Test of a cross-sectional model. International Journal of Behavioral Nutrition and Physical Activity 16, 1–17 (2019).

44. Beckers, D., Karssen, L. T., Vink, J. M., Burk, W. J. & Larsen, J. K. Food parenting practices and children’s weight outcomes: A systematic review of prospective studies. Appetite 158, 105010 (2021).

45. Rodgers, R. F. et al. Maternal feeding practices predict weight gain and obesogenic eating behaviors in young children: A prospective study. International Journal of Behavioral Nutrition and Physical Activity 10, 1–10 (2013).

46. Cardel, M. et al. Parental feeding practices and socioeconomic status are associated with child adiposity in a multi-ethnic sample of children. Appetite 58, 347–353 (2012).

47. Gross, R. S., Mendelsohn, A. L., Fierman, A. H., Racine, A. D. & Messito, M. J. Food insecurity and obesogenic maternal infant feeding styles and practices in low-income families. Pediatrics 130, 254– 261 (2012).

48. Barrett, M., Spires, M. & Vogel, C. The Healthy Start scheme in England “is a lifeline for families but many are missing out”: a rapid qualitative analysis. BMC Med 22, 1–15 (2024).

49. Caspi, C. E., Sorensen, G., Subramanian, S. V. & Kawachi, I. The local food environment and diet: A systematic review. Health Place 18, 1172 (2012).

50. Paquet, C. et al. The moderating role of food cue sensitivity in the behavioral response of children to their neighborhood food environment: A cross-sectional study. International Journal of Behavioral Nutrition and Physical Activity 14, 1–12 (2017).

51. Hoenink, J. C. et al. The moderating role of eating behaviour traits in the association between exposure to hot food takeaway outlets and body fatness. International Journal of Obesity 2023 47:6 47, 496–504 (2023).

52. Vogel, C. et al. Examination of how food environment and psychological factors interact in their relationship with dietary behaviours: Test of a cross-sectional model. International Journal of Behavioral Nutrition and Physical Activity 16, 1–17 (2019).

53. Rogers, N. T. et al. Health impacts of takeaway management zones around schools in six different local authorities across England: a public health modelling study using PRIMEtime. BMC Medicine 2024 22:1 22, 1–13 (2024).

54. Taheem, R., Woods-Townsend, K., Lawrence, W., Baird, J. & Godfrey, K. M. How do local authority plans to tackle obesity reflect systems thinking? Perspect Public Health 143, 324–336 (2023).

55. Stansfield, J., South, J. & Mapplethorpe, T. What are the elements of a whole system approach to community-centred public health? A qualitative study with public health leaders in England’s local authority areas. BMJ Open 10, e036044 (2020).

56. Bates, B., et al. National Diet and Nutrition Survey Rolling Programme Years 9 to 11 (2016/2017 to 2018/2019). https://assets.publishing.service.gov.uk/government/uploads/system/uploads/attachment_data/file/943114/NDNS_UK_Y9-11_report.pdf (2020).

57. Public Health England. NDNS: Diet and physical activity – a follow-up study during COVID-19 - GOV.UK. https://www.gov.uk/government/statistics/ndns-diet-and-physical-activity-a-follow-up-study-during-covid-19 (2021).

58. Cobiac, L. J. et al. Impact of the UK soft drinks industry levy on health and health inequalities in children and adolescents in England: An interrupted time series analysis and population health modelling study. PLoS Med 21, e1004371 (2024).

59. Water Only Schools | Healthy Schools. https://www.london.gov.uk/what-we-do/health/healthy-schools-london/awards/node/2847.

60. Hall, J. et al. Reflections on co-producing an obesity-prevention toolkit for Islamic Religious Settings: a qualitative process evaluation. International Journal of Behavioral Nutrition and Physical Activity 21, 1–12 (2024).

61. Food Insecurity Tracking | Food Foundation. https://foodfoundation.org.uk/initiatives/food-insecurity-tracking#tabs/Round-12.

