## Supplemental Files for "The impact of deprivation and neighbourhood food environments on home food environments, parental feeding practices, and child eating behaviours, food preferences and BMI: The Family Food Experience Study-London"

**Understanding family food norms and obesity-related behaviours in the context of deprivation and the wider food system: The Family Food Experience Study London**

Supplementary materials

**Findings from FFES-L pilot study and feasibility study**

A pilot feasibility study of the sampling strategy, recruitment, study materials, fieldworker training, the question and measurement order of the survey, flow and length was carried out in May to June 2021 to test various aspects of the survey. It also provided the opportunity to test a remote interviewing model over the phone (whilst in-home interviewing was not permitted due to COVID-19 restrictions). The target of the survey pilot and feasibility was to recruit two schools, one in the most and one in the least deprived neighbourhoods (defined using IDACI) and a goal to recruit on average 23 families per school. Recruitment at the school-level was challenging but met. However, at the family-level, recruitment was deemed unsatisfactory with only 11/46 target families recruited, and all the families were from the least deprived wards. The pilot also revealed that the initial interview slightly exceeded the 1-hour target (71.8 minutes long) and took considerably longer for families for whom English was not their primary language (83.8 vs 62.2 minutes for families whose primary language was English). NatCen interviewers highlighted that caregiver enjoyed the interview and that completion by phone was acceptable.

**Table S1.** Summary of targets and result of the feasibility study for FFES-L

|  | **Target for pilot study** | **Contacted** | **Consented into study/achieved** |
| --- | --- | --- | --- |
| **Schools** | 1 from least deprived ward | 10 | 1 |
|  | 1 from most deprived ward | 10 | 1 |
| **Families** | 23 from least deprived ward | n/a | 11 |
|  | 23 from most deprived ward | n/a | 0 |
| **Interview** | 60 min |  | 71.8 min (average) |

Interviewers however raised concerns about the comprehension of the consent section and several survey items. The terminology used was too complicated and not immediately understood by parents. This was particularly problematic for parents whose primary language was not English. To overcome recruitment challenges, it was agreed that schools would additionally be contacted via the local council. NatCen also developed an official accreditation logo that schools could display on their website to publicise their involvement with the FFES- L research study. To increase the clarity and comprehensibility of the interview survey, an informal translator was encouraged to support caregivers whose primary language was not English, and the wording on the survey and study material was simplified to reduce the reading age. This also shortened the time needed to complete the interview. Recruitment for the main data collection began following these changes

**Fig S1.** Flowchart of recruitment and sample for the Family Food Experiences Study - London survey (n=739 families)

**R1: Schools approached**

Total n=152

- Higher deprivation wards n=75 schools
- Lower deprivation wards: n=77 schools

**Excluded Schools**

**No response (n=100)**

- Higher deprivation: n=48

- Lower deprivation: n=52

**Refused (n=24)**

- Higher deprivation: n=10/75

- Lower deprivation: n=14/77

**R2: Schools approached^1^**

Total n=136

**Excluded Schools**

**No response (n=87)**

**Refused (n=17)**

**Home interviews (n=739)**

- Face-to-face (n=225)
- Telephone (n=514)

**Schools consented into study (n=60)**

R1 Total n=28

- Higher deprivation wards n=17 schools
- Lower deprivation wards: n=11 schools

R2 Total n=32

**Active recruitment schools**

R1 Total n=28 schools approached

- Responsive higher deprivation ward schools (n=11)
- Responsive lower deprivation ward schools (n=9)

R2 Total n=32 schools approached

- Responsive schools (n=10)

**Parental opt-in from schools**

R1 Total n=412

- Parent opt-in from higher deprivation ward schools (n=233)
- Parent opt-in from lower deprivation ward schools (n=261)
- Additional opt-ins^4^ (n=56)

R2 Total n=588

- Responsive schools (n=470)
- Additional sample issuing^5^ (n=83)

**SCHOOL RECRUITMENT**

**FAMILY RECRUITMENT**

^1:^In Round 2, the remaining 136 schools in the 4 boroughs were selected for the second round of recruitment. These schools fall in the middle of the deprivation scores so have not been split into most or least deprived wards.

^2^ No school name provided but eligible address and primary school aged children in their household.

^3^ Ineligible cases were those who had only 1 primary school aged child in their household who was in Year 6 during the recruitment period prior to September 2021, because this child would be in secondary school during the data collection period (from September 2021 to May 2021) and would therefore not be eligible for the study.

^4^ Additional cases who have opted-in during the second round of recruitment but have come from schools that agreed to take part in R1.

^5^ Additional inclusion of 83 cases who had previously been removed from issuing because they live outside the 4 selected London Boroughs but still live in London and have provided enough information to be included

^6^ Deadwood refers to an ineligible, vacant or non-residential address.

***Abbreviations:*** *R1= Round 1 from 19^th^ April – 4^th^ June 2021; R2= Round 2 from 31^st^ August to 31st December 2021.*

**Excluded Schools**

**No response (n=30)**

- Higher deprivation: n=6

- Lower deprivation: n=2

**Refused (n=24)**

**Excluded**

Refusal (n-156)

No contact (n=60)

Unproductive (n=33)

Deadwood^6^ (n=12)

**Excluded**

Unknown^2^ (n=122)

Ineligible^3^ (n=35)

**Table S2.** Representativeness of FFES-L study sample compared to wider Local Authority data on ethnicity, child weight status and IMD

| **Characteristics** | **Sample** | **Local Authority** |
| --- | --- | --- |
|  | **Mean (SD) or N (%)** | **Mean or %** |
| **Croydon (sample n=208)** |  |  |
| **Ethnicity** |  |  |
| White | 96 (**46.2**) | 48.4 |
| Mixed or multiple ethnic groups | 42 (**20.2**) | 7.6 |
| Asian or Asian British | 23 (**11.1**) | 17.5 |
| Black, Black British, Caribbean, or African | 43 (**20.7**) | 22.6 |
| Other ethnic group | 4 (**1.9**) | 3.9 |
| **Child weight status – Reception (n=50)** |  |  |
| Underweight | 2 **(4.0)** | 1.9 |
| Healthy Weight | 40 **(80.0)** | 77.7 |
| Overweight/obesity | 8 **(16.0)** | 20.3 |
| **Child weight status – Year 6 (n=11)** |  |  |
| Underweight | 1 **(9.1)** | 1.3 |
| Healthy Weight | 8 **(72.7)** | 58.7 |
| Overweight/obesity | 2 **(18.2)** | 39.9 |
| **IMD – average rank score** | 13863.57 | 18371.23 |
| **Greenwich (sample n=147)** |  |  |
| **Ethnicity** |  |  |
| White | 80 (**54.4**) | 55.7 |
| Mixed or multiple ethnic groups | 31 (**21.1**) | 6 |
| Asian or Asian British | 13 (**8.8**) | 13.2 |
| Black, Black British, Caribbean, or African | 20 (**13.6**) | 21 |
| Other ethnic group | 3 (**2.0**) | 4.2 |
| **Child weight status – Reception (n=35)** |  |  |
| Underweight | 2 **(5.7)** | 1.2 |
| Healthy Weight | 24 **(68.6)** | 77.0 |
| Overweight/obesity | 9 **(25.7)** | 21.7 |
| **Child weight status – Year 6 (n=11)** |  |  |
| Underweight | 0 **(0.0)** | 1.6 |
| Healthy Weight | 4 **(36.4)** | 57.1 |
| Overweight/obesity | 7 **(63.6)** | 41.2 |
| **IMD – average rank score** | 12943.27 | 20383.77 |
| **Haringey (sample n=180)** |  |  |
| **Ethnicity, No. (%)** |  |  |
| White | 80 (**44.4**) | 57 |
| Mixed or multiple ethnic groups | 31 (**17.2**) | 7 |
| Asian or Asian British | 7 (**3.9**) | 8.7 |
| Black, Black British, Caribbean, or African | 54 (**30.0**) | 17.6 |
| Other ethnic group | 8 (**4.4**) | 9.7 |
| **Child weight status – Reception (n=29)** |  |  |
| Underweight | 3 **(10.3)** | 2.0 |
| Healthy Weight | 19 **(65.5)** | 78.9 |
| Overweight/obesity | 7 **(24.1)** | 19.7 |
| **Child weight status – Year 6 (n=7)** |  |  |
| Underweight | 0 **(0.0)** | 1.5 |
| Healthy Weight | 4 **(57.1)** | 59.9 |
| Overweight/obesity | 3 **(42.9)** | 38.6 |
| **IMD – average rank score** | 8332.96 | 21887.62 |
| **Southwark (sample n=193)** |  |  |
| **Ethnicity, No. (%)** |  |  |
| White | 85 (**44.0**) | 51.4 |
| Mixed or multiple ethnic groups | 32 (**16.6**) | 7.2 |
| Asian or Asian British | 15 (**7.8**) | 9.9 |
| Black, Black British, Caribbean, or African | 56 (**29.0**) | 25.1 |
| Other ethnic group | 5 (**2.6**) | 6.3 |
| **Child weight status – Reception (n=34)** |  |  |
| Underweight | 2 **(5.9)** | 1.2 |
| Healthy Weight | 24 **(70.6)** | 76.8 |
| Overweight/obesity | 8 **(23.5)** | 21.7 |
| **Child weight status – Year 6 (n=14)** |  |  |
| Underweight | 1 **(7.1)** | 1.3 |
| Healthy Weight | 5 **(35.7)** | 57.2 |
| Overweight/obesity | 8 **(57.1)** | 41.5 |
| **IMD – average rank score** | 11021.35 | 21247.36 |

Ethnicity: from 2021 Census data

IMD: English Indices of Deprivation 2019, which provides a set of relative measures of deprivation for small areas (Lower-layer Super Output Areas) across England, based on seven domains of deprivation. The domains were combined using the following weights to produce the overall Index of Multiple Deprivation: Income Deprivation (22.5%); Employment Deprivation (22.5%); Education, Skills and Training Deprivation (13.5%); Health Deprivation and Disability (13.5%); Crime (9.3%); Barriers to Housing and Services (9.3%); Living Environment Deprivation (9.3%). The IMD ranks every small area in England from 1 (most deprived area) to 32,844 (least deprived area).

Child weight status: From National Child Measurement Programme, 2021/2022 school year

**Table S3.** Definitions and rating for food outlets. Scores for food outlets ranged from 10 to -10, where 10 represented the healthiest food outlets and -10 represented the least healthy outlets. * Definitions and scoring informed by Moayyed et al, 2017

| Food outlet classification | Outlet Description | Food Outlet Rating |
| --- | --- | --- |
| Fast-food chain | Outlet mainly engaged in the sale of meals that are ready for immediate consumption; table service not provided; meal can be eaten on site or taken away; the food outlet is a franchise/chain store with food being sold in specialised packaging. E.g. McDonalds, KFC, Subway, Burger King, Domino’s, Gregg’s | -10 |
| Petrol station | Outlet based in at a petrol/service station mainly selling a limited food items (e.g. Shell, Service station) | -10 |
| Specialty food store, extra foods | Outlet mainly engaged in the sale of foods than can be defined under extra food, such as ice-creams, donuts, confectionery, waffles, cakes E.g. ShakeAway, Chatime, Sprinkles Gelato | -8 |
| Independent take-away | Outlet mainly engaged in the sale of meals/snacks that are ready for immediate consumption; table service not provided; meals can be eaten on site or taken away; the outlet is not a franchise. E.g. kebab, fish & chips, burger, fried chicken and pizza outlets | -8 |
| Bargain/ Pound/ Variety Stores | Outlet not specialising in food sales; often have a range of packaged food items for sale at a reduced price E.g. Poundland, The Range, B&M Bargains | -5 |
| Convenience store | Outlet mainly engaged in the sale of a limited line of groceries generally includes milk, bread and canned and packaged foods. E.g. Corner shops, One Stop, McColls | -5 |
| Restaurant | Outlet mainly engaged in the sale of meals/snacks for consumption on the premises; table service provided; may sell alcohol with food; may provide takeaway services. E.g. franchise restaurants and non-franchise restaurants such as Harvester, Nandos, Pizza Express | 0 |
| Café/Coffee Shop | Outlet mainly engaged in the sale of light meals and hot drinks such as tea and coffee. E.g. Costa, Starbucks, sandwich shops | 0 |
| Delicatessen | Mainly engaged in the sale of specialty packaged or fresh products such as cured meats and sausage, pickled vegetables, dips, bread and olives; may also provide dine in meals. | 0 |
| Bakery | Outlet mainly serve bread, biscuits, cakes, pastries, or other flour products with or without packaging. May be independent or chain bakeries (not including Gregg’s) | 0 |
| Small supermarket | Outlet mainly engaged in the sale of groceries (fresh foods, canned and packaged food, dry goods) of non-specialised (conventional) food lines. Usually have 4 or fewer checkouts and typically belong to a large supermarket chain. E.g. Tesco’s Express, Sainsburys Local, Co-op | 5 |
| Large supermarket | Outlet mainly engaged in the sale of groceries (fresh foods, canned and packaged foods, dry goods) of non-specialised (conventional) food lines. May contain a butcher or baker. Usually have 5 or more checkouts. E.g. Sainsburys, Tesco’s, Waitrose, Lidl, ALDI | 5 |
| Butchers | Mainly engaged in the sale of fresh meat; including wholesale stores with direct to public sales. | 9 |
| Fishmongers | Mainly engaged in the sale of fresh seafood; including wholesale stores with direct to public sales and takeaway stores also providing a range of fresh seafood. | 9 |
| Greengrocers/ local produce | Mainly engaged in the sale of fresh fruit and vegetables, including wholesale stores with direct to public sales | 10 |

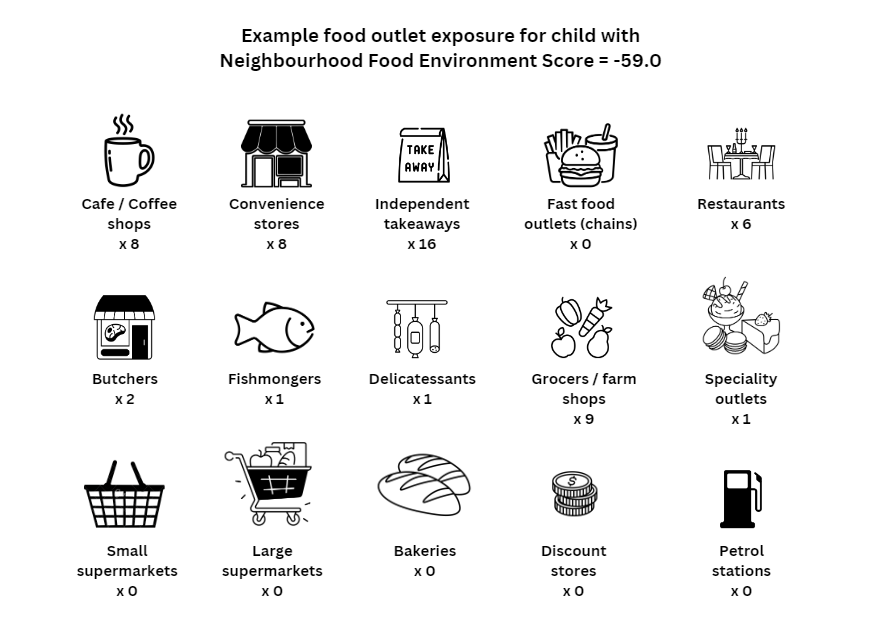

**Figure S2.** Example of food outlet exposure for a child with a neighbourhood food environment score of -59.0.

**Measures – Supplementary information**

**Table S4.** Additional information on measures used to assess parental feeding practices, child eating behaviours, food preferences and the neighbourhood food environment in FFES-L

**Parental feeding practices**

‘Instrumental feeding’ measures caregivers’ use of food as a contingency for healthy food consumption or good behaviour (4 items; e.g., *‘I use puddings as a bribe to get my child to eat his/her main course’; α = 0.70*) (Wardle et al., 2002). ‘Emotional feeding’ measures caregivers’ use of food to manage or control a child’s negative emotions (5 items; e.g. *‘I give my child something to eat to make him/her feel better when s/he is feeling upset’; α = 0.85*) (Wardle et al., 2002). ‘Pressure to eat’ measures caregivers’ attempts to coerce the child to eat more (5 items; e.g. *‘My child should always eat all of the food I give him/her’; α = 0.63*) (Birch et al., 2001). ‘Restriction’ was measured using a scale specifically designed to capture parental tendency to limit a child’s access to, and portion sizes of, sugary and high fat foods (4 items, e.g. ‘*I limit my child’s access to sugary foods’; α = 0.81*) (Cooke, 2008). ‘Parent control’ examines caregiver control over what their child eats at meals and snacks, and when and where they eat (5 items; e.g. *‘I let my child decide when s/he would like to have his/her meal’; α = 0.30*) (Wardle et al., 2002). ‘Encouragement to eat’ assesses caregivers’ use of positive reinforcement to encourage their child to eat food (such as praise for trying a new food), particularly healthy foods (5 items; e.g. *‘I encourage my child to eat a wide variety of foods’; α = 0.53*) (Wardle et al., 2002). ‘Monitoring’ assesses the extent to which caregivers keep track of their child’s high fat/sugary food consumption while in their own or others’ care (3 items; e.g. *‘I keep track of the high fat foods that my child eats’; α = 0.66*) (Birch et al., 2001). ‘Modelling’ assesses the extent to which caregivers model healthy eating for their children (4 items; e.g. *‘I model healthy eating for my child by eating healthy foods myself’; α = 0.73*) (Musher-Eizenman & Holub, 2007). ‘Covert restriction’ measures the extent to which parents restrict their child’s access to foods, supposedly without their child knowing (4 items; e.g. *‘I avoid buying unhealthy foods and bringing them into the house’; α = 0.67*) (Ogden et al., 2006). Mealtime structure measures the extent to which parents provide structure around feeding interactions (3 items; e.g. *‘My child has a set mealtime and snack routine’; α = 0.36*). A mean score was calculated for a scale if responses were available for most items within a scale (e.g., If participants had completed ≥2/3 items for monitoring, ≥3/4 items for modelling, restriction, covert restriction, and >3/5 items for remaining scales).

**Child eating behaviours**

Food Responsiveness (FR) measures a child’s drive to eat in response to external food cues (5 items e.g. ‘Given the choice, my child would eat most of the time’; α = 0.79). Enjoyment of Food (EF) assesses a child’s subjective pleasure from eating (4 items, e.g. ‘My child loves food’; α = 0.84). Emotional Overeating (EOE; 4 items, e.g. ‘My child eats more when worried’; α = 0.76) and Emotional Undereating (EUE; 4 items, e.g. ‘My child eats less when s/he is tired’; α = 0.75) assess the extent to which a child eats (more or less) in response to emotional stressors. Desire to drink (DD) measures a child’s wanting for beverages (3 items, e.g. ‘My child is always asking for a drink’; α = 0.77). Satiety Responsiveness (SR) measures a child’s sensitivity to internal cues of ‘fullness’ (5 items e.g. ‘My child gets full up easily’; α = 0.75). Slowness in Eating (SE) refers to the speed at which a child eats (4 items, e.g. ‘My child eats slowly’; α = 0.78). Food Fussiness (FF) examines a child’s pickiness about the flavour and texture of foods they are willing to eat, and their willingness to try new/unfamiliar foods (6 items, e.g. ‘My child refuses new foods at first’; α = 0.53). A mean score was calculated for each subscale only if participants had completed the majority of items for that scale (3/4 for EOE, EUE, EF, SE, 2/3 for DD, 3/5 for FR and SR, and 4/6 for FF).

**Food preferences**

Foods were categorised into six groups: fruit (α = 0.90), vegetables (α = 0.87), protein (e.g. meat and fish; α = 0.85), dairy (e.g. cheese, yogurt, eggs; α = 0.72), starches (e.g. rice, bread and pasta; α = 0.43), and snacks (e.g. chocolate, cookies, ice cream, chips; α = 0.76).

**Neighbourhood food environment**

POI data were summarised for each child’s activity space using Ordnance Survey POI data released in December 2021 from EDINA digimap. Food outlets identified from the POI dataset were reclassified, if necessary, to align with the definitions for food outlets of interest in a previous Delphi study (**Table S3**) (Moayyed et al., 2017), using a combination of data sources from previous desk-based food outlet classification methods (Lake et al., 2012; Wilkins et al., 2019). A spatial join operation was performed in the Geographic Information System (GIS) to merge information from the child’s activity space to the POI data. This produced a list of POI locations in each child’s activity space, including the number and type of food outlets present. GIS analysis was carried out in ArcGIS Pro 3.1 and ArcGIS Online.

Using the food outlet data, a score for each child was calculated that represented the type and number of food outlets they were exposed to in their activity space. These scores represented both spatial exposure to different types of food outlets and a proxy of the healthfulness of the in-store environment based on the availability of healthy and unhealthy foods in each type of outlet. Scores were calculated by: (i) identifying the number of each type of food outlet within the child’s activity space, and (ii) multiplying the number of each food outlet by a weighting describing the relative availability of healthy and unhealthy foods within each type of food outlet. The weights were determined from a previous Delphi study which consulted 17 public health and nutrition experts (Moayyed et al., 2017). In this Delphi study experts were provided with a list of 24 types of food outlets, complete with definitions and preliminary healthiness ratings based on scientific evidence. They were invited to adjust these rating through two rounds of feedback. Scores for food outlets ranged from 10 to -10, where 10 represented the healthiest food outlets and -10 represented the least healthy outlets. The food outlets and corresponding ratings used in this study are shown in Table S3.

**References**

Moayyed, H., Kelly, B., Feng, X. & Flood, V. Evaluation of a ‘healthiness’ rating system for food outlet types in Australian residential communities. *Nutrition & Dietetics* **74**, 29–35 (2017).

Lake, A. A., Burgoine, T., Stamp, E., & Grieve, R. (2012). The foodscape: Classification and field validation of secondary data sources across urban/rural and socio-economic classifications in England. International Journal of Behavioral Nutrition and Physical Activity, 9(1), 1–12. <https://doi.org/10.1186/1479-5868-9-37/TABLES/7>

Wilkins, E., Morris, M., Radley, D., & Griffiths, C. (2019). Methods of measuring associations between the Retail Food Environment and weight status: Importance of classifications and metrics. SSM - Population Health, 8. https://doi.org/10.1016/J.SSMPH.2019.100404
